# The Chest CT Features of Coronavirus Disease 2019 (COVID-19) in China: A Meta-analysis of 19 Trials

**DOI:** 10.1101/2020.05.31.20118059

**Authors:** Haitao Yang, Yuzhu Lan, Xiujuan Yao, Sheng Lin, Baosong Xie

## Abstract

**Objective:** This study aimed to summarize the characteristics of chest CT imaging in Chinese patients with Coronavirus Disease 2019 (COVID-19) to provide reliable evidence for further guiding clinical routine.

**Methods:** PubMed, Embase and Web of Science databases were thoroughly searched to identified relevant articles involving the features of chest CT imaging in Chinese patients with COVID-19. All data were analyzed utilizing R software version i386 4.0.0. Random-effects models were employed to calculate pooled mean differences.

**Results:** 19 trials incorporating 1332 cases were included in the study. The results demonstrated that the incidence of ground-glass opacities (GGO) was 0.79, consolidation was 0.34; mixed GGO and consolidation was 0.46; air bronchogram sign was 0.41; crazy paving pattern was 0.32; interlobular septal thickening was 0.55; reticulation was 0.30; bronchial wall thickening was 0.24; vascular enlargement was 0.74. subpleural linear opacity was 0.28; intrathoracic lymph node enlargement was 0.03; pleural effusions was 0.03. The distribution in lung: the incidence of central was 0.05; peripheral was 0.74; peripheral involving central was 0.38; diffuse was 0.19; unifocal involvement was 0.09; multifocal involvement was 0.57; unilateral was 0.16; bilateral was 0.83; The incidence of lobes involved (>2) was 0.70; lobes involved (≦2) was 0.35.

**Conclusion:** GGO, vascular enlargement, interlobular septal thickening more frequently occurred in patients with COVID-19. Peripheral, bilateral, involved lobes >2 might be the features of COVID-19 in the distribution aspect. Therefore, based on the aboved features of COVID-19 in chest CT imaging, it might be a promising means for identifying COVID-19.

## Introduction

The Coronavirus disease 2019 (COVID-19) is caused by a new coronavirus (SARS-COV-2) of the Sarbe virus subgenus, a member of the orthocoronavirus subfamily [1]. The outbreak of the COVID-19 has resulted in a global pandemic. Up to May 14, 2020, a total of 4 248 389 confirmed cases have been reported in the world, with another 292 046 confirmed deaths, distributed across 216 countries[2]. Considering COVID-19 has caused a big threat to global health, WHO announced the event constituted a Public Health Emergency of International Concern (PHEIC), on December 30, 2019. Interrupting the spread of the pandemic has become an urgent problem. In the prevention and treatment of SARS-COV-2, the “four early” principles (early detection, early diagnosis, early isolation and early treatment) are particularly important. Patients infected with COVID-19 may have fever, cough, dyspnea, and muscle pain, which are nonspecific [3–6]. However, the varieties of clinical manifestations, laboratory tests and imaging tests limit to clinical diagnosis and treatment. As we all know, SARS-CoV-2 nucleic acid detection is the reference standard, but it has a high false-negative rate due to nasopharyngeal swab sampling error, which often requires repeated samples. Moreover, subclinical cases increase the difficulty of diagnosis. Some studies have shown that patients with COVID-19 may have no clinical manifestations, but can find abnormal signals in chest CT [7–9]. Imaging can be used to guide diagnosis early in the course of the disease or in asymptomatic patients. Chest CT played an important role in timely detecting lung abnormalities, allowing for early treatment. Previous studies focused on the features of CT imaging of COVID-19, whereas the results varieties of different studies. Therefore, it is urgent to conduct this meta-analysis to comprehensively summarize the characteristics of CT imaging of patients with COVID-19 to further guide clinical and scientific research through evidence-based medicine.

## Material and Methods

### 2.1 Search strategy

Relevant articles were thoroughly searched from PubMed, Embase and Web of Science databases using the following words: “2019-nCoV”, “Coronavirus”, “COVID-19”, “SARS-CoV-2”, “Chest computer tomography (CT) manifestations”, “Imaging findings”. Articles were dated up to 13 May 2020. The language was restricted to English. The identified articles with the references were also searched for extending the search. All recruited articles were performed by two researchers.

### 2.2 Study selection

All articles meeting the following criteria were identified in this study: i) all patients with COVID-19 were proved by RT-PCR; ii) all articles investigated the features of chest CT imaging with sufficient data; iii) all articles were published in English. Reviews, letters, case reports, ongoing studies and studies with insufficient data were excluded.

### 2.3 Data extraction

Two researchers extracted the data from eligible articles independently. The following data included clinical characteristics (author, published year, nation, sample size, gender, age-range, fever, cough, myalgia or fatigue, sore throat, dyspnea, diarrhea, nausea, and vomiting) and the features of chest CT (ground-glass opacities (GGO), consolidation, mixed GGO and consolidation, air bronchogram sign, crazy paving pattern, interlobular septal thickening, reticulation, bronchial wall thickening, vascular enlargement, subpleural linear opacity, intrathoracic lymph node enlargement, pleural effusions, central, peripheral, peripheral involving central, unilateral, bilateral, diffuse, unifocal involvement, multifocal involvement, number of lobes involved(>2), number of lobes involved (≤2)). The third researcher made a decision once the extracted data existed discrepancy.

### 2.4 Statistical analysis

All data were analyzed utilizing R software version i386 4.0.0. Eligible data were first transformed by one of the methods (raw, i.e. untransformed, proportions (PRAW), log transformation (PLN), logit transformation (PLOGIT), arcsine transformation (PAS), Freeman-Tukey double arcsine transformation (PFT)) to make them conform to normal distribution. Random-effects models were employed to calculate pooled mean differences due to the existed incorporate heterogeneity.

## Result

### 3.1 Literature search and clinical characteristics

Firstly, the aboved terms were used to comprehensively search from 3 databases and included 21214 articles. Secondly, 11988 articles were excluded by checking duplication. Furthermore, 9171 articles were excluded by reading abstract and title; 36 articles were excluded by reading full text. Eventually, 19 articles (1332 cases) [10–19] [7, 20–27]were recruited to perform this meta-analysis to describe the chest CT feature of COVID-19. The process of searching was shown in Figure 1, and the clinical features of the included studies were described in Table 1.

**Figure 1.**
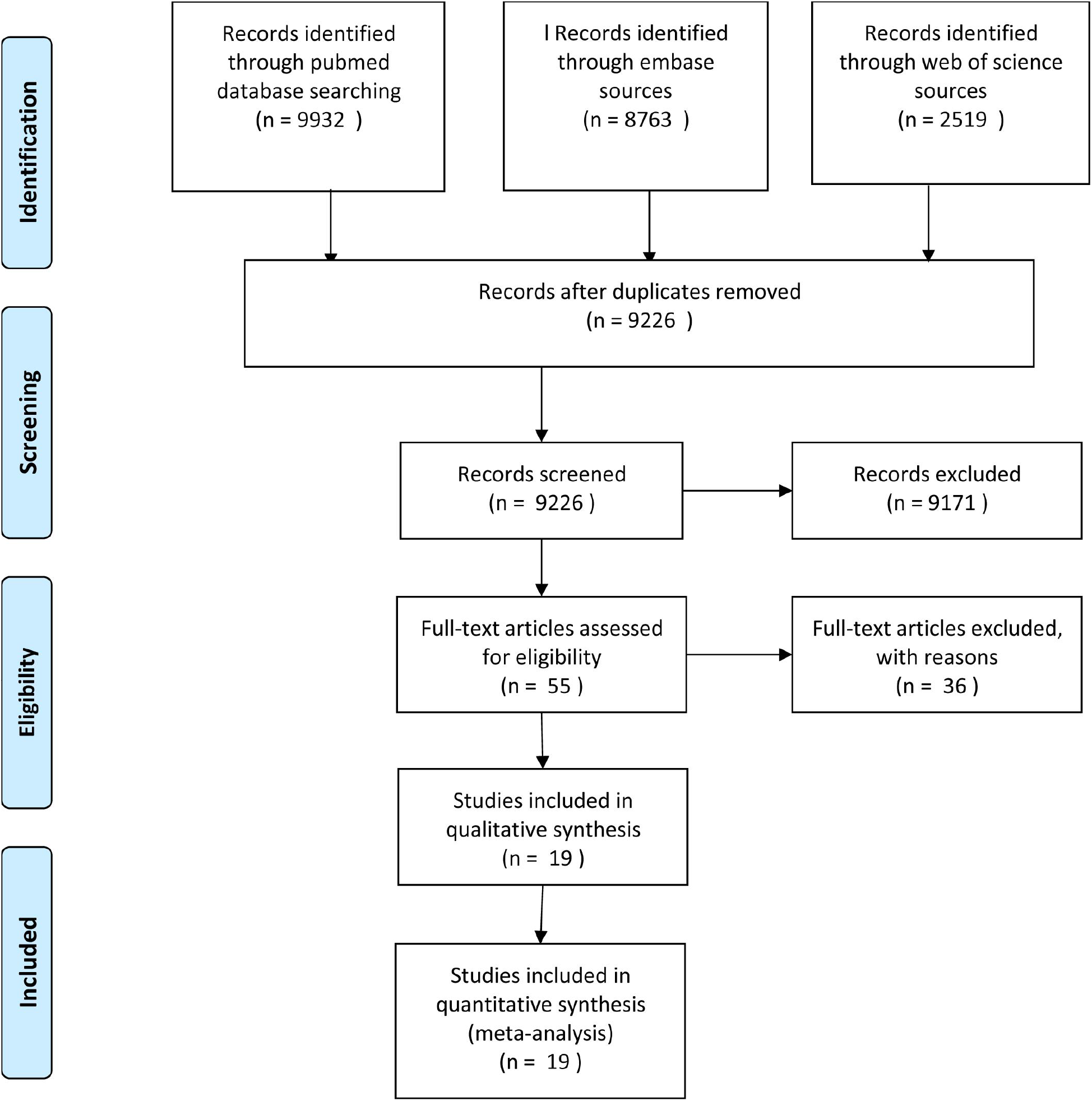
The Flow chart for study selection.

**Table 1.** the characteristics of included studies.

| Author | Publuish<br>year | Nation | Sample<br>size | Gender |  | Age range | Symptoms |  |  |  |  |  |  |  |  |
| --- | --- | --- | --- | --- | --- | --- | --- | --- | --- | --- | --- | --- | --- | --- | --- |
|  |  |  |  | male | female |  | Fever | Cough | Myalgia or<br>fatigue | Sore throat | Dyspnea | Diarrhea | Nausea and<br>vomiting | Sputum | Headache |
| Zhao W,et al | 2020 | China | 101 | 56 | 45 | 44.4(17-75) | 79 | 63 | 17 | 12 | 1 | 3 | 2 | NA | NA |
| Xu X,et al | 2020 | China | 90 | 39 | 51 | 50(18-86) | 70 | 57 | 25 | 23 | NA | 5 | 2 | NA | NA |
| Han R,et al | 2020 | China | 108 | 38 | 70 | 45(21-90) | 94 | 65 | 56 | 14 | NA | 15 | NA | NA | NA |
| Yan Li | 2020 | China | 53 | 28 | 23 | 58(26-83) | 46 | 1 | 3 | NA | NA | NA | NA | NA | NA |
| Xiong Y,et al | 2020 | China | 42 | 25 | 17 | 49.5(26-75) | 36 | 27 | 14 | NA | 8 | 10 | NA | NA | NA |
| Cheng Z, et al | 2020 | China | 11 | 8 | 3 | 50.36 | 8 | 7 | 3 | 1 | 1 | NA | NA | 3 | NA |
| Xu Y, et al | 2020 | China | 50 | 29 | 21 | 13.9(3-85) | 43 | 20 | 16 | 4 | 4 | 1 | 1 | NA | 5 |
| Yuan M, et al | 2020 | China | 27 | 12 | 15 | 60(47-69) | 21 | 16 | 3 | NA | 11 | NA | NA | NA | NA |
| Chung M, et al | 2020 | China | 21 | 13 | 8 | 51(29-77) | 14 | 9 | 6 | NA | NA | NA | 1 | NA | 3 |
| Zhang S, et al | 2020 | China | 17 | 8 | 9 | 48.6(23-74) | 12 | 9 | 13 | 1 | 1 | NA | NA | 7 | 4 |
| Pan Y, et al | 2020 | China | 63 | 33 | 30 | 44.9 | NA | NA | NA | NA | NA | NA | NA | NA | NA |
| Shi H, et al | 2020 | China | 81 | 42 | 39 | 19.5(25-81) | 59 | 48 | 7 | NA | 34 | 3 | 4 | 15 | 5 |
| Li K, et al | 2020 | China | 90 | 44 | 39 | 45.5 | 72 | 65 | 15 | 6 | 9 | 7 | NA | 15 | 9 |
| Bernheim A, et al | 2020 | China | 121 | 61 | 60 | 45.3(18-80) | 74 | 58 | NA | NA | NA | NA | NA | 20 | NA |
| Wu J, et al | 2020 | China | 80 | 38 | 42 | 44 | 61 | 58 | 13 | 9 | 7 | 7 | NA | 11 | 8 |
| Guan C,et al | 2020 | China | 53 | 25 | 28 | 42(1-86) | NA | NA | NA | NA | NA | NA | NA | NA | NA |
| Bai H, et al | 2020 | China | 219 | 119 | 100 | 44.8±14.5;<br>(4-76) | 142 | NA | NA | NA | NA | NA | NA | NA | NA |
| Song F, et al | 2020 | China | 51 | 25 | 26 | 49±16 | 49 | 24 | 16 | 3 | 7 | 5 | 3 | NA | 8 |
| Miao C, et al | 2020 | China | 54 | 28 | 26 | 45.1±13.4 | NA | NA | NA | NA | NA | NA | NA | NA | NA |

NA: not available.

### 3.2 The features of chest CT in COVID-19

#### 3.2.1 GGO, consolidation, mixed GGO, and consolation, air bronchogram sign

The results showed that the incidence of GGO was 0.79, 95% CI (0.68; 0.89) (*I^2^* = 95%, *P*<0.01) (Figure 2A), consolation was 0.34, 95% CI (0.23; 0.47) (*I^2^* = 95%, *P*<0.01) (Figure 2B), mixed GGO and consolation was 0.46, 95% CI (0.37; 0.56) (*I^2^* = 86%, *P*<0.01) (Figure 2C), air bronchogram sign was 0.41, 95% CI (0.26; 0.55) (*I^2^* = 96%, *P*<0.01) (Figure 2D).

**Figure 2.**
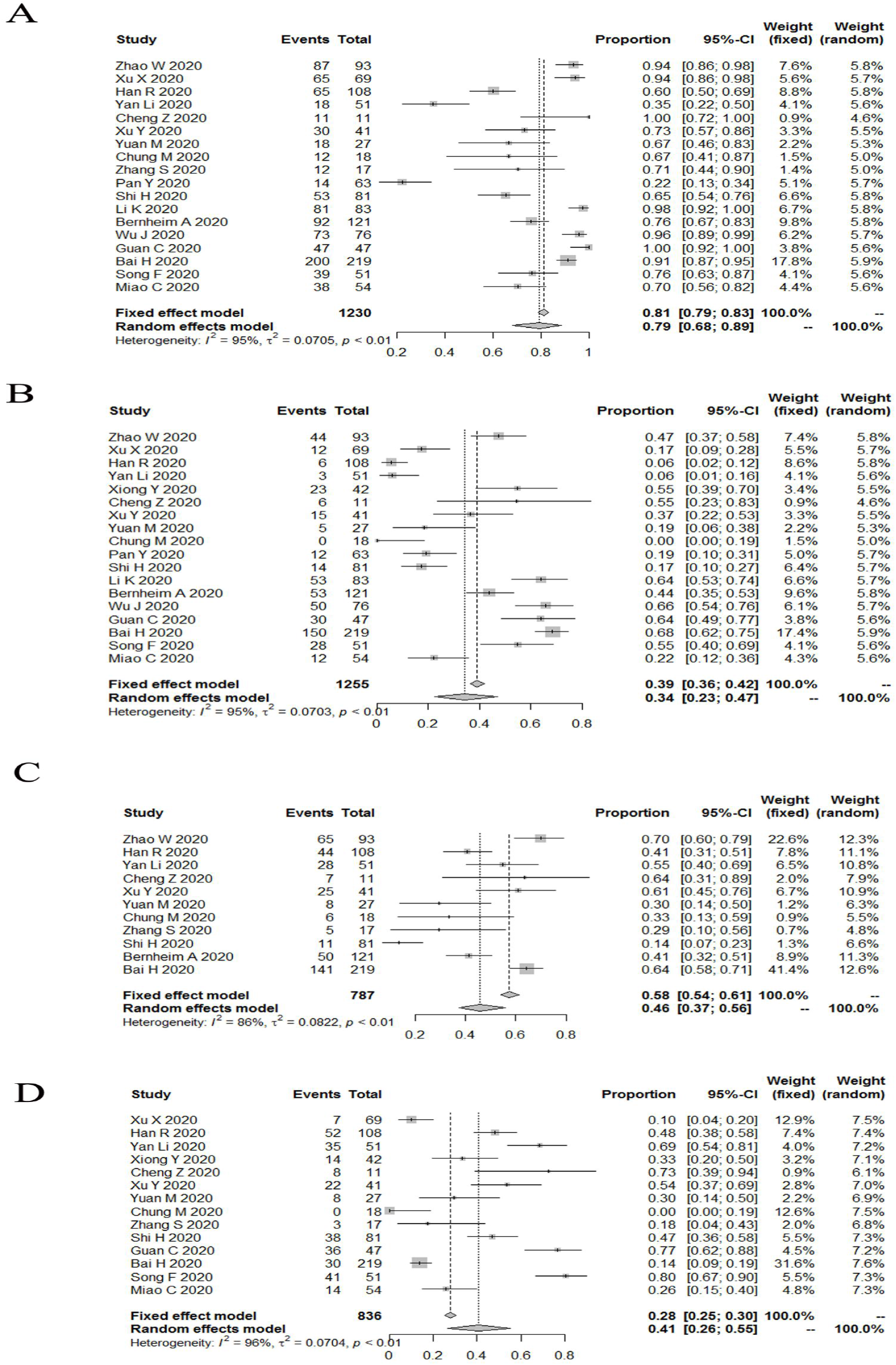
The incidence of GGO (2A), consolidation (2B), mixed GGO and consolidation (2C), air bronchogram sign (2D) in patients with COVID-19.

#### 3.2.2 The changes of pulmonary interstitial

The results reported that the incidence of crazy paving pattern was 0.32, 95% CI (0.17; 0.47) (*I^2^* = 98%, *P*<0.01) (Figure 3A); interlobular septal thickening was 0.55, 95% CI (0.42; 0.67) (*I^2^* = 84%, *P*<0.01) (Figure 3B); reticulation was 0.30, 95% CI (0.12; 0.48) (*I^2^* = 97%, *P*<0.01) (Figure 3C); bronchial wall thickening was 0.24, 95% CI (0.11; 0.40) (*I^2^* = 94%, *P*<0.01) (Figure 3D); vascular enlargement was 0.74, 95% CI (0.64; 0.86) (*I^2^* = 86%, *P*<0.01) (Figure 3E); subpleural linear opacity was 0.28, 95% CI (0.12; 0.48) (*I^2^* = 92%, *P*<0.01) (Figure 3F).

**Figure 3.**
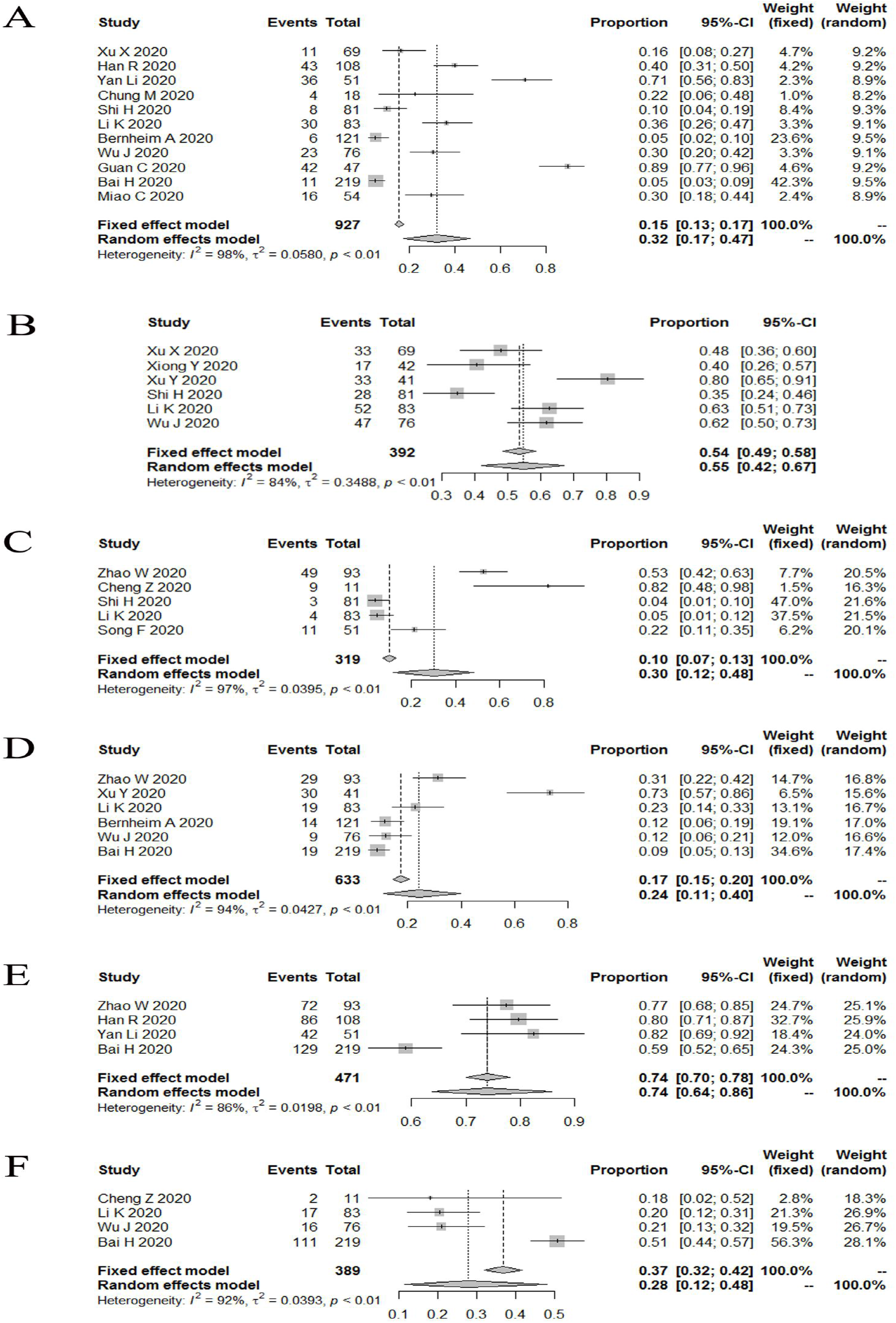
The incidence of Crazy paving pattern (3A), interlobular septal thickening (3B), reticulation (3C), bronchial wall thickening (3D), vascular enlargement (3E), subpleural linear opacity (3F) in patients with COVID-19.

#### 3.2.3 Rare signs

The results proved that the incidence of; intrathoracic lymph node enlargement was 0.03, 95% CI (0.00; 0.07) (*I^2^* = 81%, *P*<0.01) (Figure 4A); pleural effusions was 0.03, 95% CI (0.02; 0.06) (*I^2^* = 68%, *P*<0.01) (Figure 4B).

**Figure 4.**
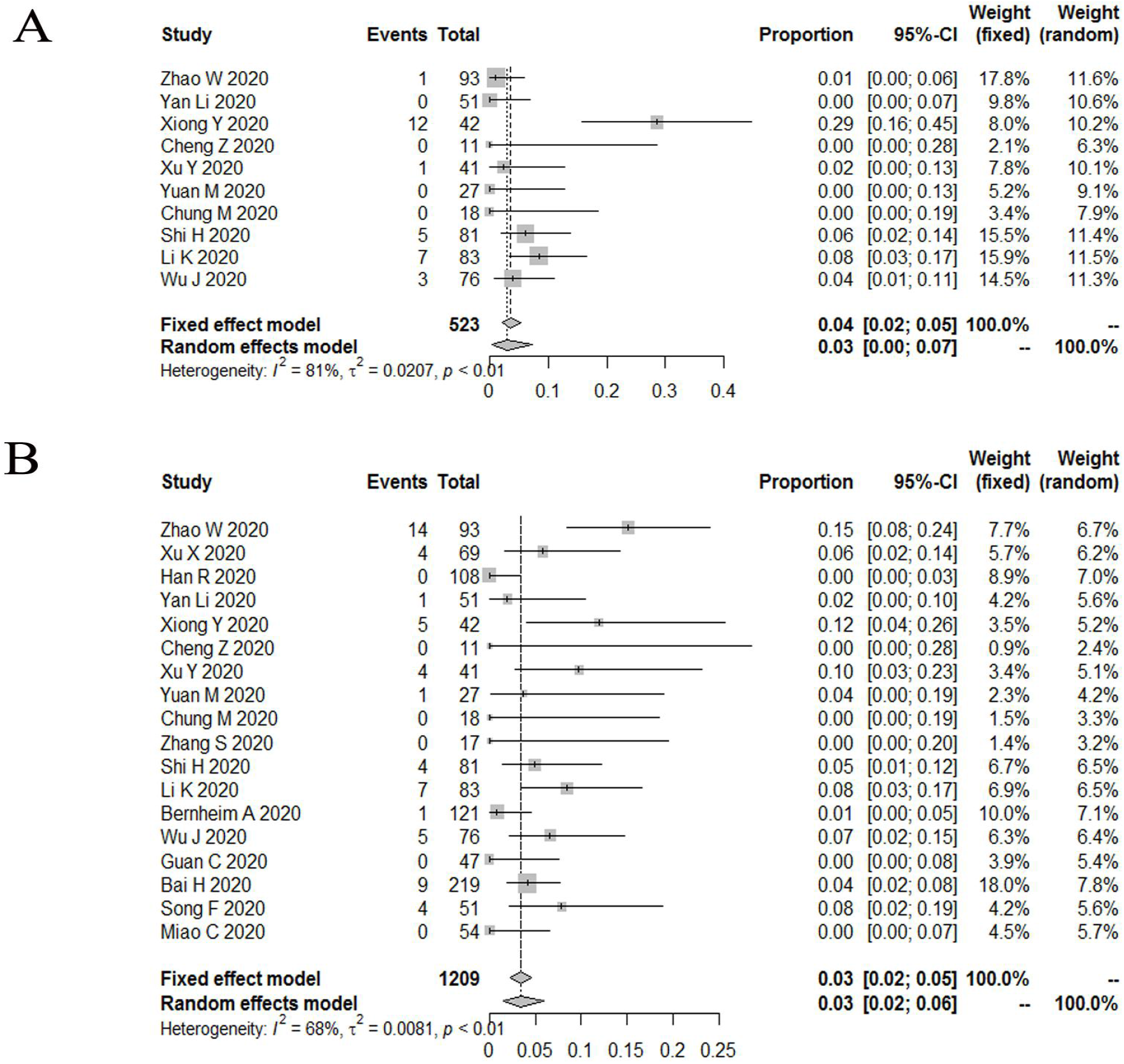
The incidence of intrathoracic lymph node enlargment (4A), pleural effusions (4B) in patients with COVID-19.

#### 3.2.4 The lesion distribution in lung

The results demonstrated that the incidence of central was 0.05, 95% CI (0.01;

0.11) (*I^2^* = 91%, *P*<0.01) (Figure 5A); peripheral was 0.74, 95% CI (0.62; 0.84) (*I^2^* = 94%, *P*<0.01) (Figure 5B); peripheral involving central was 0.38, 95% CI (0.19; 0.75) (*I^2^* = 96%, *P*<0.01) (Figure 5C); diffuse was 0.19, 95% CI (0.06; 0.32) (*I^2^* = 96%, *P*<0.01) (Figure 6A); unifocal involvement was 0.09, 95% CI (0.05; 0.14) (*I^2^* = 58%, *P* = 0.07) (Figure 6B); multifocal involvement was 0.57, 95% CI (0.48; 0.68) (*I^2^* = 80%, *P*<0.01) (Figure 6C); unilateral was 0.16, 95% CI (0.10; 0.23) (*I^2^* = 84%, *P*<0.01) (Figure 7A); bilateral was 0.83, 95% CI (0.78; 0.89) (*I^2^* = 89%, *P*<0.01) (Figure 7B); number of lobes involved(>2) was 0.70, 95% CI (0.61; 0.78) (*I^2^* = 79%, *P*<0.01) (Figure 8A); number of lobes involved(≦2) was 0.35, 95% CI (0.26; 0.44) (*I^2^* = 80%,*P*<0.01) (Figure 8B).

**Figure 5.**
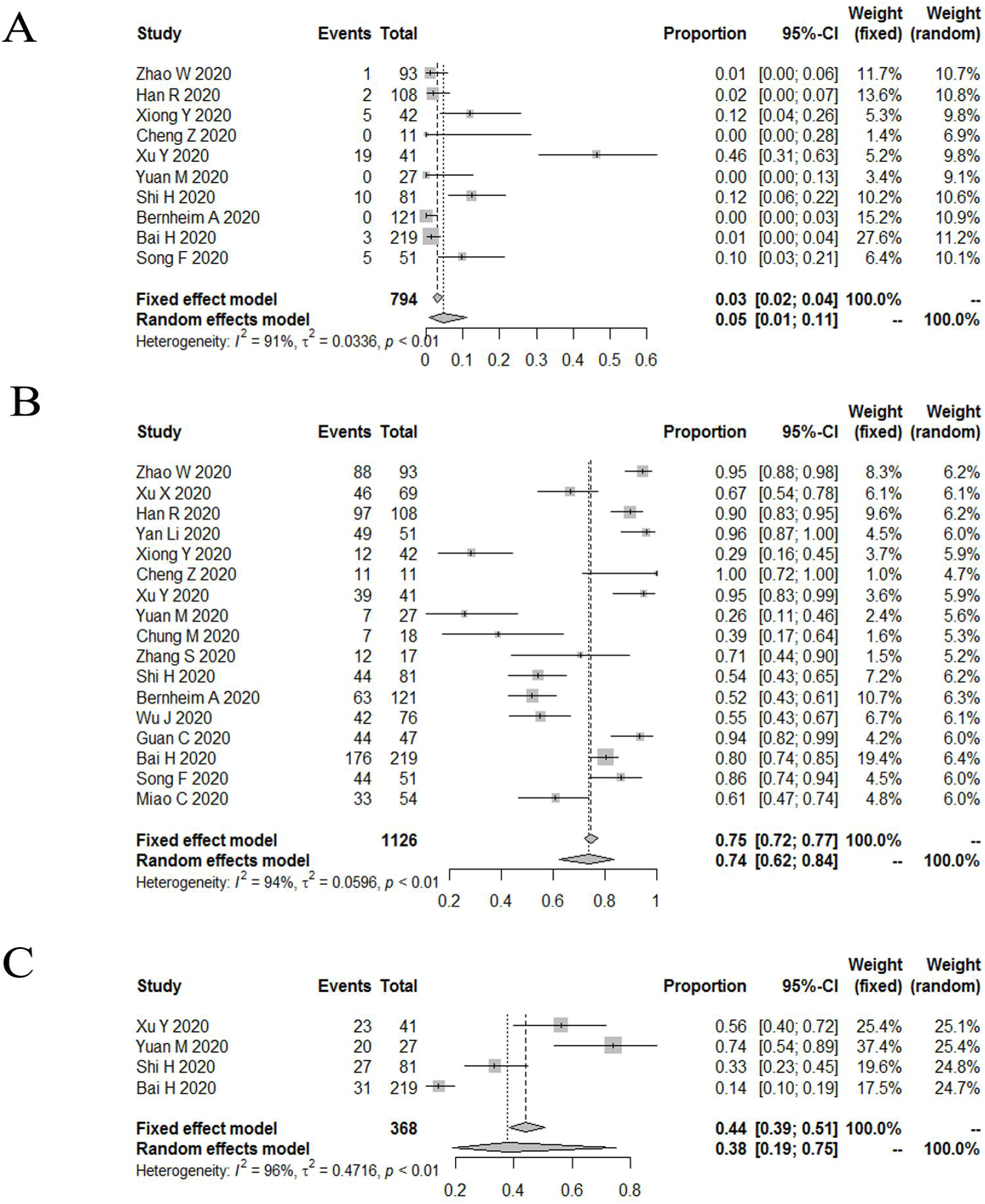
The incidence of central (5A), peripheral (5B), peripheral involving central (5C) in patients with COVID-19.

**Figure 6.**
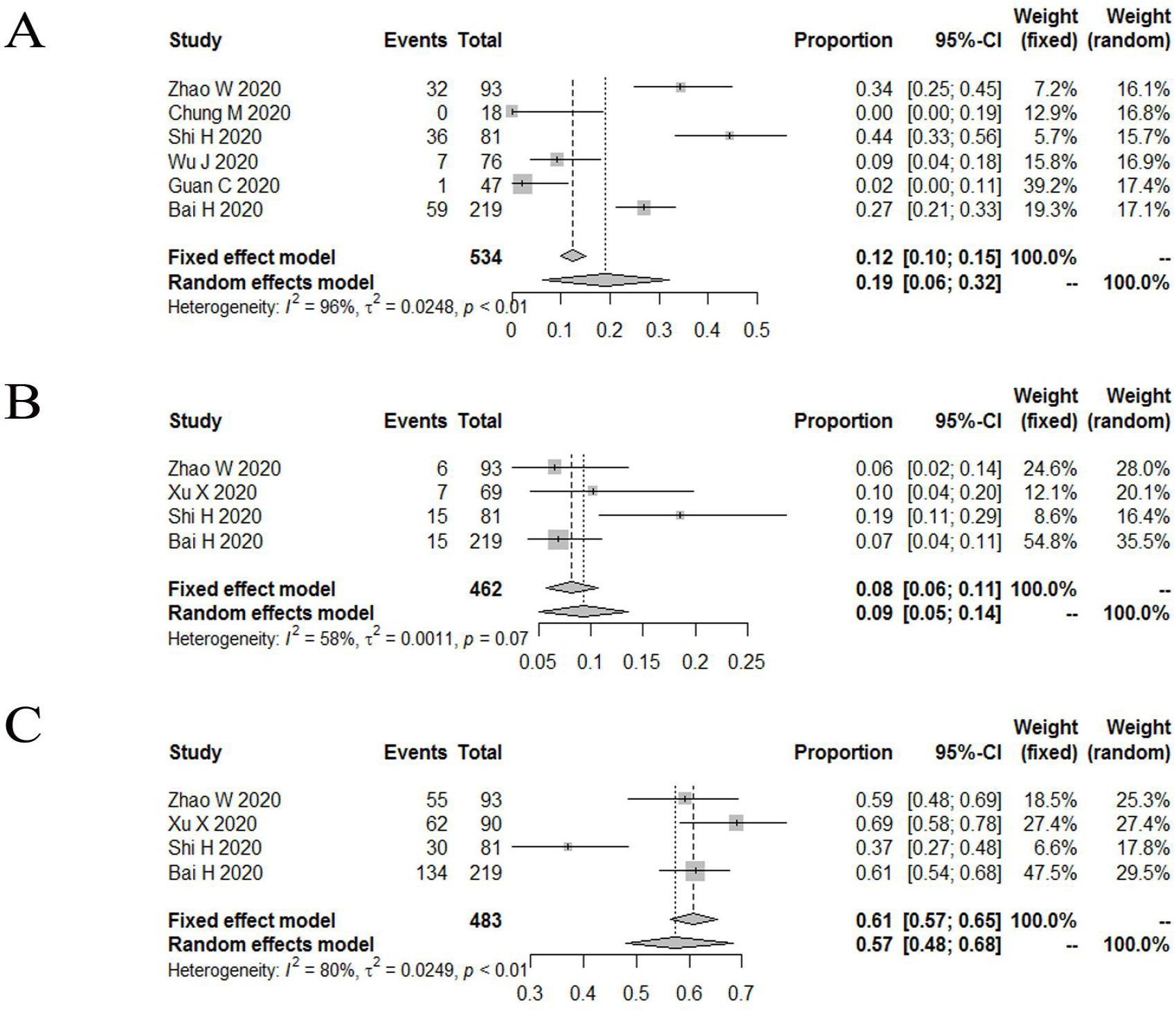
The incidence of diffuse (6A), unifocal involvement (6B), multifocal involvement (6C) in patients with COVID-19.

**Figure 7.**
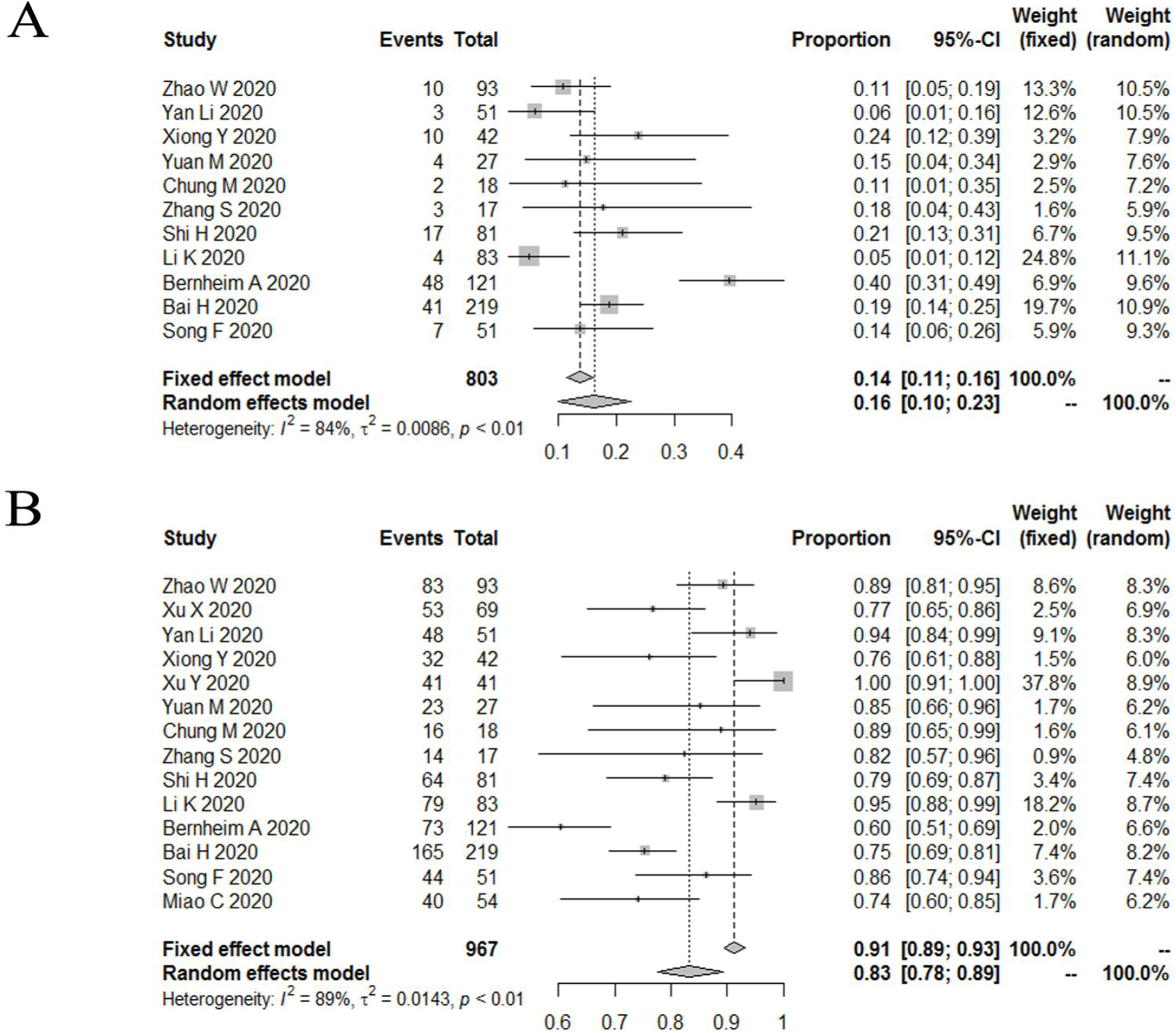
The incidence of unilateral (7A), bilateral (7B) in patients with COVID-19.

**Figure 8.**
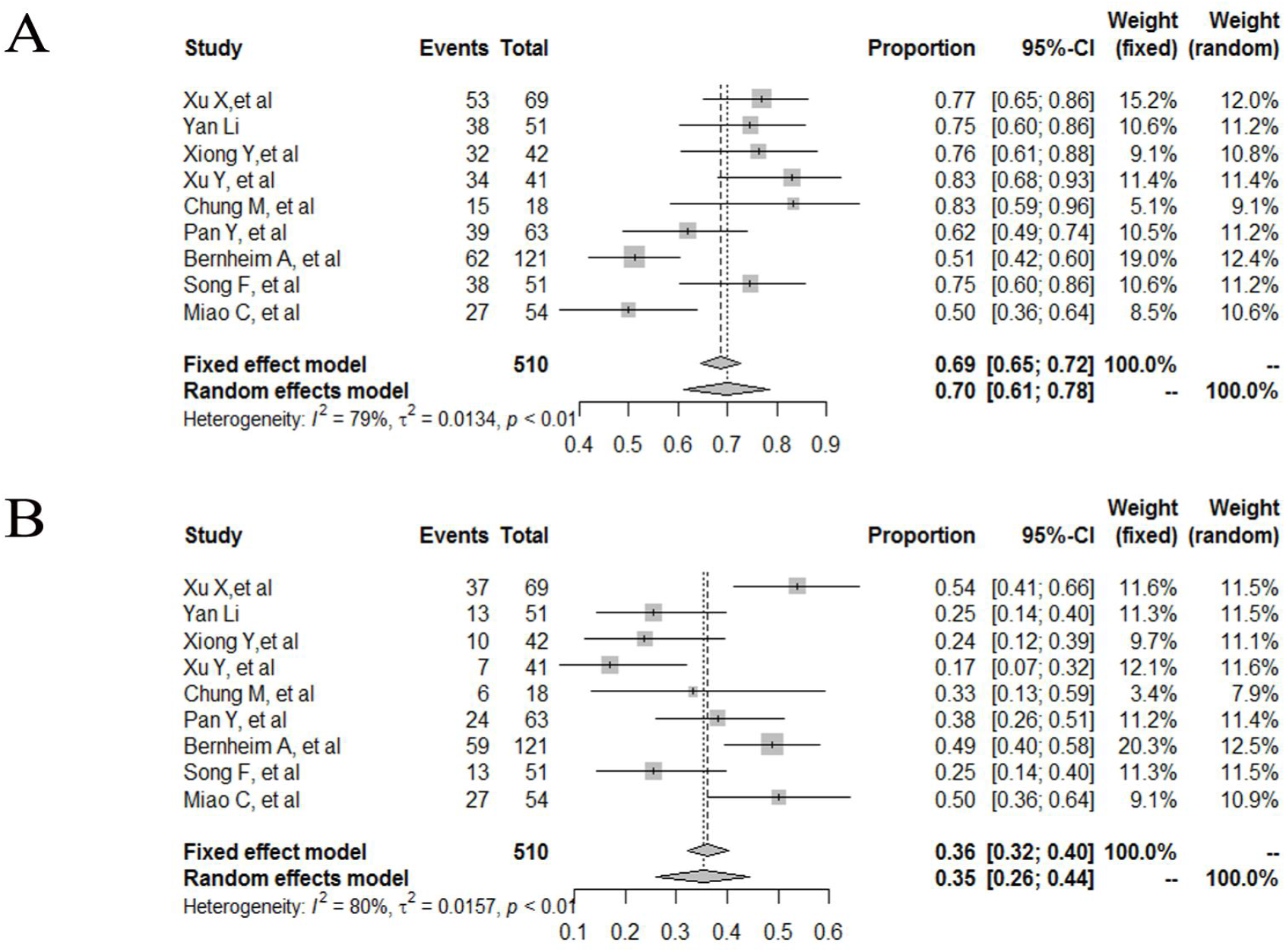
The incidence of number of lobes involved (>2) (8A), number of lobes involved (≦2) (8B) in patients with COVID-19.

## Discussion

This novel coronavirus disease is known as COVID-19 by the world health organization [28, 29]. Early detection, early diagnosis, early isolation and early treatment principle are important to control this disease. The RT-qPCR method, as the gold standard for detection of pathogens, is characterized by high sensitivity and specificity. But it requires laboratory-level facility, reliable power supply, expensive equipment and trained personnel to properly conduct RT-qPCR tests, which limits its application to some extent [30]. In addition to the nucleic test, CT also can be helpful to diagnose COVID-19. The diagnosis of viral pneumonia based on radiologic features by radiologists as one of the diagnostic criteria for COVID-19 according to the diagnosis and treatment program (6th version) published by the National Health Commission of the People’s Republic of China [31]. High-resolution CT is highly sensitive to detect lung abnormalities, which is quite helpful for early diagnosis of the disease that can trigger early treatment and facilitates to contain this emergency disease [32, 33]. Some articles have shown the detailed CT features of COVID-19 [34–36]. Our study may conclude some common CT imaging features in patients affected by SARS-CoV-2 pneumonia.

To the best of our knowledge, this study involving 19 trials (1332 cases) is the first meta-analysis to extensively summarize the characteristics of chest CT imaging in patients with COVID-19. Our study revealed that GGO, vascular enlargement, interlobular septal thickening more frequently occurred in patients with COVID-19. Peripheral, bilateral, involved lobes > 2 might be the features of COVID-19 in the distribution aspect. In this study, intrathoracic lymph node enlargement, pleural effusions, the lesion distribution in lung of central, unifocal, and unilateral were not frequently observed. Therefore, based on the aboved features of COVID-19 in chest CT imaging, it might be a promising means for identifying COVID-19.

The CT patterns of viral pneumonia are related to the pathogenesis of viral infection. Viruses from the same family (e.g Coronaviridae) have similar pathogenesis [37]. The SARS-CoV-2 belongs to the family Coronaviridae, which includes other viruses like Severe Acute Respiratory Syndrome (SARS) and Middle East Respiratory Syndrome (MERS) [28]. Some investigations[35, 38, 39] have shown that SARS-CoV-2 pneumonia CT findings were partially similar to other viral pneumonia, like SARS, MERS and H7N9 pneumonia[40–42]. The pathological changes included thickening of the basement membrane of the alveoli capillary, edema of the alveoli septum, pulmonary hyaline membrane formation, inflammatory cell infiltration and inflammatory edema, pulmonary interstitial hyperplasia and fibrosis, apoptosis of alveolar epithelium cells [43, 44]. Based on the image characteristic of SARS-CoV-2 pneumonia which had been reported in some articles [43, 45–47], the pathological of GGOs may be the thickening of alveolar wall, collapse of alveolar cavity, reduction of air content in alveolar cavity and inflammatory cells infiltration or a combination of these features. We estimated the pathological mechanism of COVID-19 includes bronchoalveolar destruction and damage to lung parenchyma near the bronchioles in the early stages [45]. In the late stage, diffuse alveolar injury and acute fibrous and organic pneumonia can be observed[48]. This pathological pattern is same as imaging pattern like GGO at first and then consolidation dense consolidative lesions, early in the disease. With the progression of the disease, lesions often turn more linear fashion with predilection for the lung periphery (and somewhat with a “crazy” paving pattern or emergence of a “reverse halo” sign).

We also observed some interstitial changes in patients with COVID-19. Hitherto, some autopsy cases have revealed the pathological features of COVID-19[49–52]. The pathological changes of pulmonary fibrosis injury in COVID-19 include extensive diffuse alveolar injury with bilateral edema, protein or fibrin exudation, and diffuse reactive proliferation of type II pneumocytes. It was even can be observed that interstitial fibroblast proliferation caused alveolar septa to thicken, forming hyaline membranes consistent with fibrosis. These pathological changes may be due to a disruption of the ACE/ACE2 (angiotensin-converting enzyme/ angiotensin-converting enzyme 2) balance[53–58], which were presented as the crazy paving pattern, interlobular septal thickening, bronchial wall thickening on chest CT.

Lymphadenopathy and pleural effusions are atypical imaging features in COVID-19 patients. Severe/critical patients showed more lymph node enlargement, and pleural effusion[7, 59]. Li et al [60] reported that lymphadenopathy and pleural effusions were poor prognostic indicators according to his logistic regression models in SARS-CoV-2 pneumonia. Similarly, lymphadenopathy and pleural effusions are important predictors of an unfavorable outcome in patients infected with MERS-CoV or avian influenza H5N1[61–63]. However, articles on the above two imaging changes are less, and more evidence is needed to verify these conclusions.

Pulmonary lesions were most commonly in the peripheral, which was related to ACE2. ACE2 has been established as a functional receptor for SARS-CoV, which plays a crucial role in the pathogenesis of COVID-19[64]. ACE2 was abundantly expressed on the surface of alveolar type II pneumocytes and the capillary endothelial cells[65], where are the targets of viral entry and replication. As the virus invades, type II pneumocytes and capillary endothelial cells are constantly destroyed, which may explain why most lesions are located peripherally.

Nevertheless, we also encountered some limitations: i) eligible studies were retrospective studies; ii) large heterogeneity among included articles might affect the reliability and stability of results we analyzed to some extent.

## Conclusion

GGO, vascular enlargement, interlobular septal thickening, more frequently occurred in patients with COVID-19. Peripheral, bilateral, involved lobes > 2 might be the features of COVID-19 in the distribution aspect. Therefore, based on the above features of COVID-19 in chest CT imaging, it might be a promising means for identifying COVID-19.

## Data Availability

I vertified all data referred to in the manuscript were availabled.

## Acknowledge

None.

## Conflict of interest

None.

## Funding

None.

## Notes

### Competing Interest Statement

The authors have declared no competing interest.

## References

[1]. Xu, X., et al., Evolution of the novel coronavirus from the ongoing Wuhan outbreak and modeling of its spike protein for risk of human transmission. Sci China Life Sci, 2020. 63(3): p. 457–460.

[2]. Coronavirus disease (COVID-19) Situation Report – 115. World Health Organization, 2020.

[3]. Ai, T., et al., Correlation of Chest CT and RT-PCR Testing in Coronavirus Disease 2019 (COVID-19) in China: A Report of 1014 Cases. Radiology, 2020: p. 200642.

[4]. Huang, C., et al., Clinical features of patients infected with 2019 novel coronavirus in Wuhan, China. Lancet, 2020. 395(10223): p. 497–506.

[5]. Yang, X., et al., Clinical course and outcomes of critically ill patients with SARS-CoV-2 pneumonia in Wuhan, China: a single-centered, retrospective, observational study. Lancet Respir Med, 2020. 8(5): p. 475–481.

[6]. Wang, D., et al., Clinical Characteristics of 138 Hospitalized Patients With 2019 Novel Coronavirus-Infected Pneumonia in Wuhan, China. JAMA, 2020.

[7]. Shi, H., et al., Radiological findings from 81 patients with COVID-19 pneumonia in Wuhan, China: a descriptive study. The Lancet Infectious Diseases, 2020. 20(4): p. 425–434.

[8]. Fang, Y., et al., Sensitivity of Chest CT for COVID-19: Comparison to RT-PCR. Radiology, 2020: p. 200432.

[9]. Xie, X., et al., Chest CT for Typical 2019-nCoV Pneumonia: Relationship to Negative RT-PCR Testing. Radiology, 2020: p. 200343.

[10]. Zhao, W., et al., Relation Between Chest CT Findings and Clinical Conditions of Coronavirus Disease (COVID-19) Pneumonia: A Multicenter Study. AJR Am J Roentgenol, 2020. 214(5): p. 1072–1077.

[11]. Xu, X., et al., Imaging and clinical features of patients with 2019 novel coronavirus SARS-CoV-2. European Journal of Nuclear Medicine and Molecular Imaging, 2020. 47(5): p. 1275–1280.

[12]. Han, R., et al., Early Clinical and CT Manifestations of Coronavirus Disease 2019 (COVID-19) Pneumonia. AJR Am J Roentgenol, 2020: p. 1–6.

[13]. Li, Y. and L. Xia, Coronavirus Disease 2019 (COVID-19): Role of Chest CT in Diagnosis and Management. AJR Am J Roentgenol, 2020. 214(6): p. 1280–1286.

[14]. Xiong, Y., et al., Clinical and High-Resolution CT Features of the COVID-19 Infection: Comparison of the Initial and Follow-up Changes. Invest Radiol, 2020. 55(6): p. 332–339.

[15]. Cheng, Z., et al., Clinical Features and Chest CT Manifestations of Coronavirus Disease 2019 (COVID-19) in a Single-Center Study in Shanghai, China. AJR Am J Roentgenol, 2020: p. 1–6.

[16]. Xu, Y.H., et al., Clinical and computed tomographic imaging features of novel coronavirus pneumonia caused by SARS-CoV-2. J Infect, 2020. 80(4): p. 394–400.

[17]. Yuan, M., et al., Association of radiologic findings with mortality of patients infected with 2019 novel coronavirus in Wuhan, China. PLOS ONE, 2020. 15(3): p. e0230548.

[18]. Chung, M., et al., CT Imaging Features of 2019 Novel Coronavirus (2019-nCoV). Radiology, 2020. 295(1): p. 202–207.

[19]. Simin Zhang, H.L.S.H., High-resolution CT features of 17 cases of Corona Virus Disease 2019 in Sichuan province, China. European respiratory JOURNAL, 2020.

[20]. Pan, Y., et al., Initial CT findings and temporal changes in patients with the novel coronavirus pneumonia (2019-nCoV): a study of 63 patients in Wuhan, China. European Radiology, 2020.

[21]. Wu, J., et al., Chest CT Findings in Patients With Coronavirus Disease 2019 and Its Relationship With Clinical Features. Investigative Radiology, 2020. 55(5): p. 257–261.

[22]. Guan, C.S., et al., Imaging Features of Coronavirus disease 2019 (COVID-19): Evaluation on Thin-Section CT. Academic Radiology, 2020. 27(5): p. 609–613.

[23]. Song, F., et al., Emerging 2019 Novel Coronavirus (2019-nCoV) Pneumonia. Radiology, 2020. 295(1): p. 210–217.

[24]. Li, K., et al., The Clinical and Chest CT Features Associated With Severe and Critical COVID-19 Pneumonia. Invest Radiol, 2020. 55(6): p. 327–331.

[25]. Bernheim, A., et al., Chest CT Findings in Coronavirus Disease-19 (COVID-19): Relationship to Duration of Infection. Radiology, 2020. 295(3): p. 200463.

[26]. Bai, H.X., et al., Performance of radiologists in differentiating COVID-19 from viral pneumonia on chest CT. Radiology, 2020: p. 200823.

[27]. Miao, C., et al., Early chest computed tomography to diagnose COVID-19 from suspected patients: A multicenter retrospective study. Am J Emerg Med, 2020.

[28]. Zhu, N., et al., A Novel Coronavirus from Patients with Pneumonia in China, 2019. New England Journal of Medicine, 2020. 382(8): p. 727–733.

[29]. https://www.who.int/emergencies/diseases/novel-coronavirus-2019/technical-guidance/naming-the-coronavirus-disease-(covid-2019)-and-the-virus-that-causes-it. World Health Organization, 2020.

[30]. Nayak, S., et al., Point-of-Care Diagnostics: Recent Developments in a Connected Age. Anal Chem, 2017. 89(1): p. 102–123.

[31]. National Health Commission of the People’s Republic of China website. Diagnosis and treatment of novel coronavirus infection (trial version 6). www.nhc.gov.cn/yzygj/s7653p/202002/8334a832

[32]. Paul, N.S., et al., Radiologic pattern of disease in patients with severe acute respiratory syndrome: the Toronto experience. Radiographics, 2004. 24(2): p. 553–63.

[33]. Chung, M., et al., CT Imaging Features of 2019 Novel Coronavirus (2019-nCoV). Radiology, 2020. 295(1): p. 202–207.

[34]. Huang, C., et al., Clinical features of patients infected with 2019 novel coronavirus in Wuhan, China. Lancet, 2020. 395(10223): p. 497–506.

[35]. Lei, J., et al., CT Imaging of the 2019 Novel Coronavirus (2019-nCoV) Pneumonia. Radiology, 2020. 295(1): p. 18.

[36]. Chung, M., et al., CT Imaging Features of 2019 Novel Coronavirus (2019-nCoV). Radiology, 2020. 295(1): p. 202–207.

[37]. Koo HJ, L.S.C.J., Radiographic and CT Features of Viral Pneumonia1. Radiographics, 2018.

[38]. Shi H, H.X.J.N., Radiological findings from 81 patients with COVID-19 pneumonia in Wuhan, China: a descriptive study. Lancet, 2020.

[39]. Song, F., et al., Emerging 2019 Novel Coronavirus (2019-nCoV) Pneumonia. Radiology, 2020. 295(1): p. 210–217.

[40]. Wang, Q., et al., Emerging H7N9 influenza A (novel reassortant avian-origin) pneumonia: radiologic findings. Radiology, 2013. 268(3): p. 882–9.

[41]. Wong KT, A.G.H.D., Thin-Section CT of Severe Acute Respiratory Syndrome: Evaluation of 73 Patients Exposed to or with the Disease1. Radiology, 2003.

[42]. Ajlan, A.M., et al., Middle East respiratory syndrome coronavirus (MERS-CoV) infection: chest CT findings. AJR Am J Roentgenol, 2014. 203(4): p. 782–7.

[43]. Nicholls, J.M., et al., Lung pathology of fatal severe acute respiratory syndrome. Lancet, 2003. 361(9371): p. 1773–8.

[44]. Lang, Z.W., et al., [A clinicopathological study on 3 cases of severe acute respiratory syndrome]. Zhonghua Bing Li Xue Za Zhi, 2003. 32(3): p. 201–4.

[45]. Xu, Z., et al., Pathological findings of COVID-19 associated with acute respiratory distress syndrome. Lancet Respir Med, 2020. 8(4): p. 420–422.

[46]. Ding, Y., et al., The clinical pathology of severe acute respiratory syndrome (SARS): a report from China. J Pathol, 2003. 200(3): p. 282–9.

[47]. Lang, Z.W., et al., [A clinicopathological study on 3 cases of severe acute respiratory syndrome]. Zhonghua Bing Li Xue Za Zhi, 2003. 32(3): p. 201–4.

[48]. Ding, Y., et al., The clinical pathology of severe acute respiratory syndrome (SARS): a report from China. J Pathol, 2003. 200(3): p. 282–9.

[49]. Barton, L.M., et al., COVID-19 Autopsies, Oklahoma, USA. Am J Clin Pathol, 2020. 153(6): p. 725–733.

[50]. Tian, S., et al., Pathological study of the 2019 novel coronavirus disease (COVID-19) through postmortem core biopsies. Mod Pathol, 2020.

[51]. Xu, Z., et al., Pathological findings of COVID-19 associated with acute respiratory distress syndrome. Lancet Respir Med, 2020. 8(4): p. 420–422.

[52]. Tian, S., et al., Pulmonary Pathology of Early-Phase 2019 Novel Coronavirus (COVID-19) Pneumonia in Two Patients With Lung Cancer. J Thorac Oncol, 2020. 15(5): p. 700–704.

[53]. Tikellis, C. and M.C. Thomas, Angiotensin-Converting Enzyme 2 (ACE2) Is a Key Modulator of the Renin Angiotensin System in Health and Disease. Int J Pept, 2012. 2012: p. 256294.

[54]. Hamming, I., et al., The emerging role of ACE2 in physiology and disease. J Pathol, 2007. 212(1): p. 1–11.

[55]. Kuba, K., et al., A crucial role of angiotensin converting enzyme 2 (ACE2) in SARS coronavirus-induced lung injury. Nat Med, 2005. 11(8): p. 875–9.

[56]. Imai, Y., et al., Angiotensin-converting enzyme 2 protects from severe acute lung failure. Nature, 2005. 436(7047): p. 112–6.

[57]. Li, Y., et al., Angiotensin-converting enzyme 2 prevents lipopolysaccharide-induced rat acute lung injury via suppressing the ERK1/2 and NF-kappaB signaling pathways. Sci Rep, 2016. 6: p. 27911.

[58]. Gaur, P., et al., Regulation, signalling and functions of hormonal peptides in pulmonary vascular remodelling during hypoxia. Endocrine, 2018. 59(3): p. 466–480.

[59]. Li, K., et al., The Clinical and Chest CT Features Associated With Severe and Critical COVID-19 Pneumonia. Invest Radiol, 2020. 55(6): p. 327–331.

[60]. Li, X., et al., Comparison of chest CT findings between COVID-19 pneumonia and other types of viral pneumonia: a two-center retrospective study. Eur Radiol, 2020.

[61]. Das, K.M., et al., CT correlation with outcomes in 15 patients with acute Middle East respiratory syndrome coronavirus. AJR Am J Roentgenol, 2015. 204(4): p. 736–42.

[62]. Das, K.M., et al., Acute Middle East Respiratory Syndrome Coronavirus: Temporal Lung Changes Observed on the Chest Radiographs of 55 Patients. AJR Am J Roentgenol, 2015. 205(3): p. W267–74.

[63]. Qureshi, N.R., et al., The radiologic manifestations of H5N1 avian influenza. J Thorac Imaging, 2006. 21(4): p. 259–64.

[64]. Hamming, I., et al., Tissue distribution of ACE2 protein, the functional receptor for SARS coronavirus. A first step in understanding SARS pathogenesis. J Pathol, 2004. 203(2): p. 631–7.

[65]. Jia, H.P., et al., ACE2 receptor expression and severe acute respiratory syndrome coronavirus infection depend on differentiation of human airway epithelia. J Virol, 2005. 79(23): p. 14614–21.

